# Fit for growing old? Financial protection before and after Indonesia’s national health insurance scheme – a repeated cross-section study

**DOI:** 10.1101/2024.03.05.24303817

**Authors:** Gindo Tampubolon

## Abstract

The world is ageing with unprecedented momentum, and large global south nations are ageing at higher speed than their northern peers. They have grown old while they have not grown rich, straining their health systems’ ability to deliver financial protection. This work aimed to assess whether Indonesia’s health insurance scheme, seven years on, has delivered equal protection for families with older members (over 60 years) as for other families.

**Methods:** Before-and-after observation study is designed to estimate how much difference the Scheme made to probabilities of catastrophic payment and financial impoverishment for the two family types. As in recent assessments, two national socioeconomic surveys were used (2013, 2021). Two level observations came from 622,125 families residing in 514 districts across the archipelago. Financial protection indicators against catastrophic payment and impoverishment were constructed following recent works. I estimated two level probit models, then plotted marginal probabilities of financial protection. A sensitivity analysis was conducted with the standard financial protection indicator.

**Finding:** After the Scheme, financial hardship for all family types has reduced by 19%. But families with older members (compared to other families) have an additional 0.7% risk of incurring catastrophic payment or financial impoverishment. And social and spatial inequalities in health persist.

**Discussion:** While the Scheme has markedly improved financial protection for all, families with older members remain at higher risk of being unprotected. The global south can prepare for an ageing world by monitoring financial protection and its social determinants and systematically distinguishing families with older members.

## Introduction

Indonesia’s health system has been transformed by the launch of the national health insurance scheme or *Jaminan Kesehatan Nasional* in 2014 – the same year similar schemes were launched in Vietnam and the US. The Indonesian scheme is a universal scheme that aims to provide access to quality health care for more than 270 million citizens by 2019, making it the largest single payer system worldwide.^1,2^ Before the Scheme, Indonesia had a fragmented and unequal health system, with different types of insurance plans for different segments of the population, such as civil servants or formal sector workers. That left many with no health insurance at all. They had to pay themselves for health services, especially those in the informal sector and rural areas.^3^ This often led to health inequity and financial hardship.

Thanks to the Scheme, Indonesia has made notable progress in expanding health coverage and improving health outcomes.^2,4,5^ But the Scheme faces many challenges including insufficient funding, low quality of care, inadequate human resources, and an ageing population.^4–7^ These challenges pose risks to the sustainability and effectiveness of the Scheme to deliver financial protection.

Indonesia is the third largest global south nation and is ageing rapidly, with 48 million older people aged over 60 years by 2035 (two thirds of total UK population then) – it has grown old before it has grown rich.^8,9^ The scheme was expressly aimed to provide security in old age, which resonates with financial protection being a key goal of health systems.^10^ In a bid to deliver a sustainable development goal of universal coverage, financial protection must not take second place, leaving vulnerable sections of the population exposed to catastrophic payments or financial impoverishment.^11–18^

Very little is known about financial protection before and after the Scheme, especially after the Scheme enrolled more than two thirds of the population or stabilised. Its most recent health system review has discussed catastrophic payments after the Scheme but made no systematic comparison with before the Scheme, accounting for confounding factors such as rural-urban distinction.^19^ Most important, recent health system assessments have not fully recognised the challenges of population ageing. With people-centredness and health improvement, financial protection is a key goal of health systems.^10^ For families with older members the financial goal is acutely relevant because of the increased likelihood of chronic conditions, multiple organ dysfunctions, memory deficits, mental disorders, and frailty.^20–27^ The burden of such conditions puts families under considerable pressure, leaving them financially exposed.

While ageing has emerged as an issue in Asia, it has not been prominent in Indonesia’s health system assessments.^28^ The most recent assessment of the Scheme found that it had started with a progressive financial footing, but has subsequently been deteriorating to a point where finally in 2019 it turned regressive.^4,5^ Ageing, however, has remained outside assessment. This work aims to bring it inside, specifically:

1. to compare catastrophic health payment and financial impoverishment (summarised as financial hardship) just before the Scheme and after, 2013 and 2021,
2. to compare the financial hardship experience of families with older members with other families (with younger members), in both years,
3. to assess whether foregone care is masked in common measures of financial protection.

This work contributes to the literature in explaining the specific challenges of ageing in rapidly ageing populations in the global south and to monitoring progress towards the goal of universal coverage. Last, it points to enduring social and spatial inequalities and the balance between national and local roles in decentralised governance.

## Materials and methods

Following Cheng and collaborators, I used the national social economic surveys, extended to fit a before-and-after design (2013 and 2021).^5,29^ As secondary data the University of Manchester exempted the research from ethical review. The surveys collect spending using the United Nations’ Classification of Individual Consumption According to Purpose (COICOP). The *outcomes* are catastrophic health payment, financial impoverishment, and financial hardship, all binary variables. *Catastrophic payment* is constructed following Thomas, Cylus and colleagues writing about middle and high income countries, because Indonesia is a middle income country.^16,17^ They compared different indicators or methods of construction and found the following to be the only one that produced an effective threshold of catastrophic payment. The construction proceeded as follows: total family consumption is removed of basic needs to get capacity to pay. The ratio of direct health payment to this capacity is compared against the 40% threshold – higher means catastrophic, otherwise non-catastrophic.^11^ Now, basic needs are determined nationwide as the average of consumption of food, shelter and utilities for families found between 25^th^ to 35^th^ percentiles of total consumption. (Any family with negative capacity to pay immediately incurred catastrophic payment when paying for health.)

Analogously, the construction of *financial impoverishment* starts with total family consumption. Once direct health payment is removed, what remains is capacity to pay. This is compared against the basic needs above – lower means impoverishment, otherwise no impoverishment.^17,30^ Together, catastrophic payment and financial impoverishment give a complementary picture of financial exposure – one prioritising basic needs, another prioritising direct health payment. A third outcome, *financial hardship* summarises whether the family incurred *any* catastrophic payment or financial impoverishment; this phrase is interchangeable with *financially exposed*. Finally, whether a family has foregone care due to lack of funds in the past quarter is used as the fourth outcome. This is atypical.

To explain financial protection for families with older members and for others, an extensive set of covariates is used. The set is larger than typical studies on financial protection in Asia and includes gender (female), age and its square, education (up to secondary school; high school or more), marital status (married; never or unmarried) and employment status (formal worker; informal worker or unemployed) of the family head, family size and its composition (number of children under five; five to ten; over ten), family economic status in quintiles of total consumption, ownership of air conditioning, refrigerator or car, any outpatient visit, any inpatient visit in the past quarter, (any foregone care when explaining financial protection outcomes, not when explaining foregone care), residence (rural; urban) and regions (Java and Bali; west; centre; east). Reflecting Indonesia’s decentralised governance, local fiscal capacity is included – an official statistic produced by the Ministry of Finance.^31,32^ Reflecting Indonesia’s archipelago, a map of the country’s 514 districts is used to construct the multilevel structure of the observations – an official geostatistic produced by the Central Statistics Agency. In sum, three data sources were collated: family consumption surveys, district fiscal statistics and their districts map.

To achieve the aims, two key covariates were used: whether any family member was over 60 years (compared to all younger) and whether the observations were after the scheme (compared to before). A before-and-after design was chosen, instead of difference-in- difference design, because the scheme was launched nationwide in 2014. There was no attempt to assign families into treatment (into the Scheme) and control (as usual).

To model financial protection and foregone care *multilevel probit models* were fitted, following a recent cross-sectional study of catastrophic payment in 133 countries^12^ and because the observations make up two levels: families residing in districts across the archipelago. To help interpretation I plotted the marginal probabilities of each outcome, distinguishing families with older members from those without. A sensitivity analysis followed: the alternative of total family consumption (instead of the capacity to pay) and the threshold of 10% were used for financial protection (typical indicator for the UN Sustainable Development Goals).

## Findings

The analytic sample is summarised in table one. Briefly, in the sample there are 6.9% families headed by women, families with older members make up 27%, and more than half live in rural areas (58%). Just one third of families have heads working in formal sectors. The top part shows that 26% of families report financial hardship due to health and 9% forego care even when needed. More importantly, across three indicators the percentages were (more than) halved after the Scheme, e.g. from 33% to 12% in catastrophic payment.

To gauge the relative strengths of the confounding factors driving the large reductions, multilevel probit models were fitted to give the coefficients in table two. There are 622,125 families residing in 514 districts which varies spatially (significant σ)

The coefficients for *After* are all negative and highly significant (p < 0.0001) – as expected, fewer families were financially exposed after the Scheme. Also as expected, having an older family member increases the probability of being financially exposed (highly significant). Equally importantly, the model explaining foregone care (last column) shows that after the Scheme *more* families forego care (positive and highly significant). If one has formed an expectation that financial protection is improving after the Scheme, this is unexpected. This is a first.

Briefly, in all models, families with female head have higher probabilities of experiencing financial hardship (positive and highly significant). Rural residents have higher risk of being financially exposed whereas formal workers have lower risk. Similarly, higher quintiles in the income distributions suffer lower risks. Districts with higher fiscal capacity also have lower risks of catastrophic payment and financial impoverishment. It is unclear why the other two have positive coefficients. Towards the bottom, regardless of the gender of family head, asset ownership (air conditioning, refrigerator, car) reduces the risk of financial exposure (negative and highly significant).

These coefficients cannot be read off simply as magnitudes because the marginal probability of a factor (such as having an older member) depends non-linearly on other factors in the model. I therefore calculated marginal probabilities and plotted them, picking out two key factors for each outcome: *After* (before or after the Scheme) and with or without *older members* – collected in table three and plotted in figure one. The last column shows the additional (Δ) probability of families with older member, 1% higher catastrophic payment before the Scheme and 0.7% afterward (panel A). As another example, 1.2% higher foregone care before the Scheme and 1.7% afterward (panel D). Analogously, reduction in (Δ) probability after the Scheme for each family type is shown as the last row for each panel – 19% lower catastrophic payment for families with older members and nearly 19% lower for other families (panel A). Another striking example is 4.3% *higher* foregone care for families with older members and 3.8% higher for other families (panel D).

To highlight new patterns across all indicators I put these probabilities into a trellis plot (figure 1). The probabilities of catastrophic payment reduced markedly by 19 percentage points for any type of family. But there are higher probabilities experienced by families with older members (solid line is above dashed line). These two patterns of marked reduction after the Scheme and of worse difference are consistent across all three indicators of financial protection.

**Figure 1.**
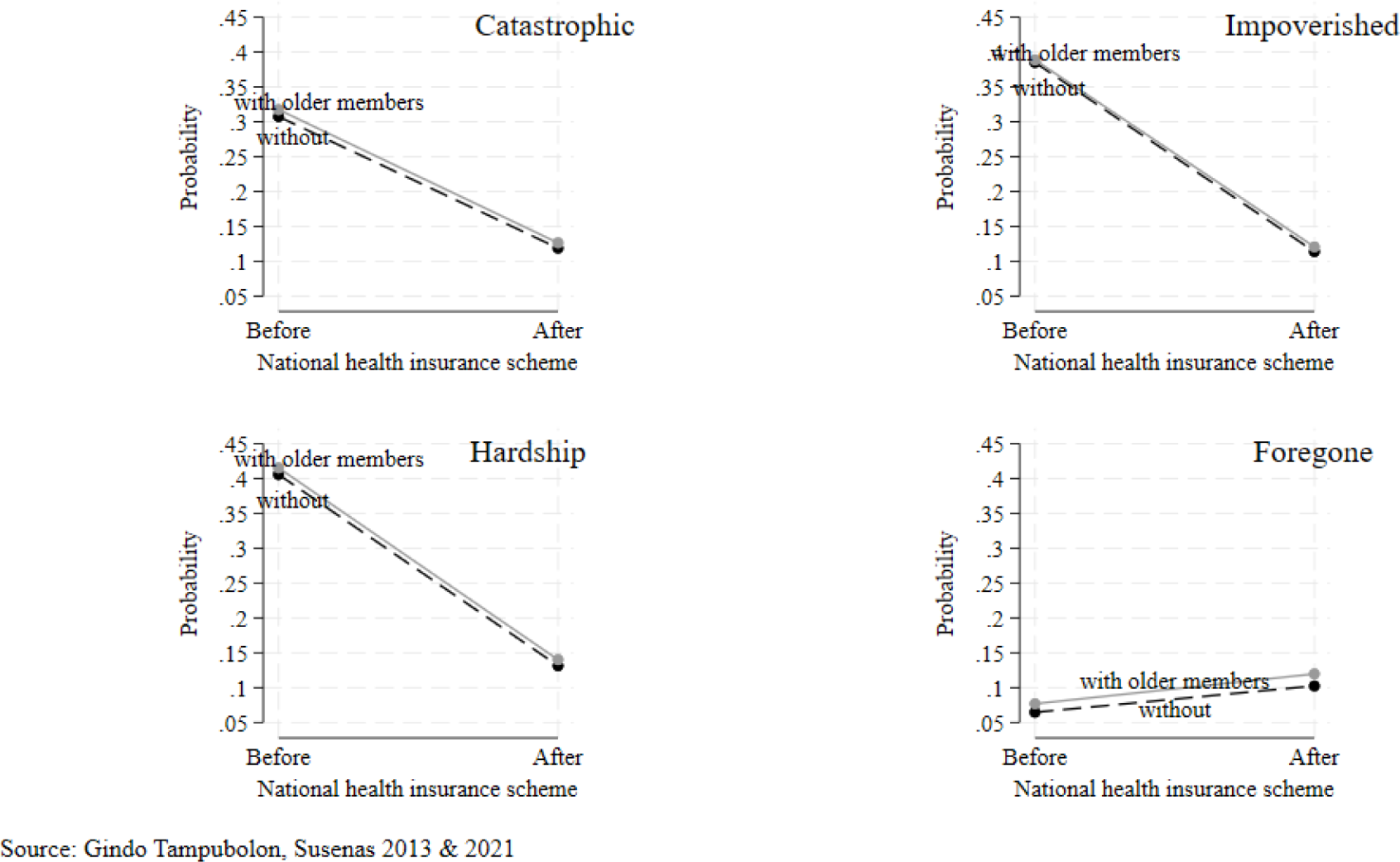
Marginal plots of probabilities of catastrophic payment, financial impoverishment, financial hardship and foregone care from multilevel probit models (table 3), highlighting before and after as well as families with older members (solid line) and without them (dashed line). Source: Susenas 2013 & 2021.

**Table 1.** Description of analytic sample of financial protection. Source: Susenas 2013, 2021

|  | Relative to national health insurance |  |  |
| --- | --- | --- | --- |
|  | Before | After | Total |
|  | N=284024 | N=339999 | N=624023 |
| Catastrophic payment |  |  |  |
| No | 191127 (67.3%) | 297810 (87.6%) | 488937 (78.4%) |
| Yes | 92897 (32.7%) | 42189 (12.4%) | 135086 (21.6%) |
| Fin. impoverished |  |  |  |
| No | 173506 (61.1%) | 299387 (88.1%) | 472893 (75.8%) |
| Yes | 110518 (38.9%) | 40612 (11.9%) | 151130 (24.2%) |
| Fin. hardship |  |  |  |
| No | 166868 (58.8%) | 294866 (86.7%) | 461734 (74.0%) |
| Yes | 117156 (41.2%) | 45133 (13.3%) | 162289 (26.0%) |
| Foregone care |  |  |  |
| No | 264716 (93.2%) | 302880 (89.1%) | 567596 (91.0%) |
| Yes | 19308 (6.8%) | 37119 (10.9%) | 56427 (9.0%) |
| Family head |  |  |  |
| Female | 18823 (6.6%) | 24246 (7.1%) | 43069 (6.9%) |
| Male | 265201 (93.4%) | 315753 (92.9%) | 580954 (93.1%) |
| With older members |  |  |  |
| No | 209355 (73.7%) | 245818 (72.3%) | 455173 (72.9%) |
| Yes | 74669 (26.3%) | 94181 (27.7%) | 168850 (27.1%) |
| Regions |  |  |  |
| Jawa Bali | 98584 (34.7%) | 111496 (32.8%) | 210080 (33.7%) |
| Sumatra | 81313 (28.6%) | 96511 (28.4%) | 177824 (28.5%) |
| Kalimantan | 28168 (9.9%) | 30786 (9.1%) | 58954 (9.4%) |
| Sulawesi | 37018 (13.0%) | 47402 (13.9%) | 84420 (13.5%) |
| Outer islands | 38941 (13.7%) | 53804 (15.8%) | 92745 (14.9%) |
| Rural |  |  |  |
| Urban | 121299 (42.7%) | 143105 (42.1%) | 264404 (42.4%) |
| Rural | 162725 (57.3%) | 196894 (57.9%) | 359619 (57.6%) |
| Log fiscal capacity | 6.48 (0.63) | 6.85 (0.72) | 6.68 (0.70) |
| Age family head | 48.2 (13.6) | 48.5 (13.5) | 48.3 (13.5) |
| Edu. family head, yr | 7.3 (4.7) | 11.4 (3.9) | 9.6 (4.7) |
| Family head employment type |  |  |  |
| Informal worker | 201144 (70.8%) | 222244 (65.4%) | 423388 (67.8%) |
| Formal worker | 82880 (29.2%) | 117755 (34.6%) | 200635 (32.2%) |
| Couple family head |  |  |  |
| Divorce/widow | 53220 (18.7%) | 65692 (19.3%) | 118912 (19.1%) |
| Couple | 230804 (81.3%) | 274307 (80.7%) | 505111 (80.9%) |
| Num. under fives | 0.3 (0.6) | 0.3 (0.6) | 0.3 (0.6) |
| Five to nine | 3.5 (1.6) | 3.4 (1.5) | 3.5 (1.6) |
| Ten or older | 3.1 (1.4) | 3.1 (1.4) | 3.1 (1.4) |
| Air conditioning |  |  |  |
| No | 271718 (95.7%) | 314752 (92.6%) | 586470 (94.0%) |
| Yes | 12306 (4.3%) | 25247 (7.4%) | 37553 (6.0%) |
| Refrigerator |  |  |  |
| No | 176870 (62.3%) | 147783 (43.5%) | 324653 (52.0%) |
| Yes | 107154 (37.7%) | 192216 (56.5%) | 299370 (48.0%) |
| Car |  |  |  |
| No | 262644 (92.5%) | 303108 (89.1%) | 565752 (90.7%) |
| Yes | 21380 (7.5%) | 36891 (10.9%) | 58271 (9.3%) |
| Any inpatient visit |  |  |  |
| No | 262094 (92.3%) | 304549 (89.6%) | 566643 (90.8%) |
| Yes | 21930 (7.7%) | 35450 (10.4%) | 57380 (9.2%) |
| Any outpatient visit |  |  |  |
| No | 187332 (66.0%) | 265142 (78.0%) | 452474 (72.5%) |
| Yes | 96692 (34.0%) | 74857 (22.0%) | 171549 (27.5%) |
| Transfer programme family |  |  |  |
| No | 278605 (98.1%) | 291087 (85.6%) | 569692 (91.3%) |
| Yes | 5419 (1.9%) | 48912 (14.4%) | 54331 (8.7%) |
| Quintiles of incomes |  |  |  |
| Poorest quintile | 56805 (20.0%) | 68000 (20.0%) | 124805 (20.0%) |
| Lower middle | 56805 (20.0%) | 68000 (20.0%) | 124805 (20.0%) |
| Middle | 56805 (20.0%) | 68000 (20.0%) | 124805 (20.0%) |
| Upper middle | 56805 (20.0%) | 68000 (20.0%) | 124805 (20.0%) |
| Richest quintile | 56804 (20.0%) | 67999 (20.0%) | 124803 (20.0%) |

**Table 2.** Coefficients and standard errors of multilevel probit models of catastrophic payment, financial impoverishment, financial hardship and foregone care. Source: Susenas 2013, 2021

|  | Catastrophic | Impoverished | Hardship | Foregone |
| --- | --- | --- | --- | --- |
| After | -1.351***<br>(0.000) | -9.343***<br>(0.000) | -3.056***<br>(0.000) | 0.258***<br>(0.000) |
| Older member | 0.065***<br>(0.000) | 0.168***<br>(0.000) | 0.100***<br>(0.000) | 0.094***<br>(0.000) |
| Foregone care | 0.302***<br>(0.000) | 0.094***<br>(0.000) | 0.058***<br>(0.000) |  |
| Sumatra | -0.299***<br>(0.000) | -0.533***<br>(0.000) | -0.256***<br>(0.000) | 0.045<br>(0.072) |
| Kalimantan | -0.213***<br>(0.000) | -0.699***<br>(0.000) | -0.319***<br>(0.000) | 0.064<br>(0.067) |
| Sulawesi | -0.229***<br>(0.000) | -0.072<br>(0.263) | -0.180***<br>(0.000) | 0.217***<br>(0.000) |
| Outer islands | -0.652***<br>(0.000) | -0.526***<br>(0.000) | -0.391***<br>(0.000) | 0.079**<br>(0.005) |
| Log fiscal capacity | -0.045***<br>(0.000) | -0.055***<br>(0.001) | 0.017**<br>(0.007) | 0.035***<br>(0.000) |
| Under five | 0.062***<br>(0.000) | -0.138***<br>(0.000) | -0.038***<br>(0.000) | 0.075***<br>(0.000) |
| Five to ten | -0.004<br>(0.374) | -0.213***<br>(0.000) | -0.105***<br>(0.000) | 0.032***<br>(0.000) |
| Eleven plus | -0.115***<br>(0.000) | -0.290***<br>(0.000) | -0.124***<br>(0.000) | -0.001<br>(0.792) |
| Lower middle q. | -0.877***<br>(0.000) | -6.481***<br>(0.000) | -1.842***<br>(0.000) | -0.043***<br>(0.000) |
| Middle quintile | -2.805***<br>(0.000) | -13.093***<br>(0.000) | -4.651***<br>(0.000) | -0.051***<br>(0.000) |
| Upper middle q. | -3.158***<br>(0.000) | -14.286***<br>(0.000) | -4.972***<br>(0.000) | -0.043***<br>(0.000) |
| Richest quintile | -2.874***<br>(0.000) | -14.513***<br>(0.000) | -4.653***<br>(0.000) | -0.047***<br>(0.000) |
| Female head | 0.227***<br>(0.000) | 0.447***<br>(0.000) | 0.219***<br>(0.000) | 0.050***<br>(0.000) |
| Age family head | 0.001<br>(0.354) | 0.002<br>(0.642) | 0.003*<br>(0.040) | -0.008***<br>(0.000) |
| Edu. family head | 0.012***<br>(0.000) | -0.005**<br>(0.009) | -0.003***<br>(0.000) | -0.003***<br>(0.000) |
| Married/couple | -0.002<br>(0.819) | -0.267***<br>(0.000) | -0.091***<br>(0.000) | 0.001<br>(0.896) |
| Formal worker | -0.111***<br>(0.000) | -0.206***<br>(0.000) | -0.134***<br>(0.000) | -0.043***<br>(0.000) |
| Rural | 0.017*<br>(0.016) | 0.124***<br>(0.000) | 0.058***<br>(0.000) | 0.008<br>(0.202) |
| Inpatient | 0.923***<br>(0.000) | 0.949***<br>(0.000) | 1.309***<br>(0.000) | -0.111***<br>(0.000) |
| Outpatient | 0.616***<br>(0.000) | 0.247***<br>(0.000) | 0.277***<br>(0.000) | -0.192***<br>(0.000) |
| Transfer programme | -0.091***<br>(0.000) | 0.019<br>(0.408) | -0.058***<br>(0.000) | 0.075***<br>(0.000) |
| Air conditioning | -0.095***<br>(0.000) | -0.723***<br>(0.000) | -0.062*<br>(0.017) | -0.131***<br>(0.000) |
| Refrigerator | -0.266***<br>(0.000) | -0.635***<br>(0.000) | -0.286***<br>(0.000) | -0.072***<br>(0.000) |
| Car | -0.258***<br>(0.000) | -0.861***<br>(0.000) | -0.254***<br>(0.000) | -0.117***<br>(0.000) |
| Constant | 1.290***<br>(0.000) | 10.686***<br>(0.000) | 3.408***<br>(0.000) | -1.762***<br>(0.000) |
| Log $\sigma$ | -2.845***<br>(0.000) | -1.759***<br>(0.000) | -3.305***<br>(0.000) | -3.183***<br>(0.000) |
| *N* | 622125 | 622125 | 622125 | 622125 |

**Table 3.** Marginal probabilities of multilevel probit models explaining four financial protection indicators using extensive covariates in table 2, picking out Before or After and With or Without older family members. Both factors are statistically significant (table two). SDG: Sustainable Development Goal.

| Pane l | Indicator | Before/<br>After | Older<br>member | Other/<br>younger | $\Delta$<br>older |
| --- | --- | --- | --- | --- | --- |
| A | Catastrophic | Before | .317 | .307 | **+0.010** |
|  |  | After | .126 | .119 | **+0.007** |
| | | $\Delta$ After | **-0.191** | **-0.188** | |
| B | Impoverished | Before | .389 | .385 | +0.004 |
|  |  | After | .121 | .114 | +0.007 |
| | | $\Delta$ After | -0.268 | -0.271 | |
| C | Hardship | Before | .415 | .406 | +0.009 |
|  |  | After | .140 | .131 | +0.009 |
| | | $\Delta$ After | -0.275 | -0.275 | |
| D | Foregone | Before | .077 | .065 | **+0.012** |
|  |  | After | .120 | .103 | **+0.017** |
| | | $\Delta$ After | **+0.043** | **+0.038** | |
| E | All inclusive | Before | .455 | .430 | **+0.025** |
|  |  | After | .258 | .239 | **+0.019** |
| | | $\Delta$ After | **-0.197** | **-0.191** | |
| F | *Sensitivity* |  |  |  |  |
|  | Catastrophic | Before | .044 | .036 | +0.008 |
|  | SDG | After | .039 | .032 | +0.007 |
| | | $\Delta$ After | -0.005 | -0.004 | |

On foregone care there has been little expectation in the literature. While the pattern of worse difference for families with older family members is retained, there is an *increase* in probability of foregone care *after* the Scheme. This upward pattern contradicts the rest, though its magnitude is the smallest. Note that 2021 is the midst of the pandemic which may explains this upward trend in foregone care – people were simply cautious to seek care.

However this trend has been observed in 2019 the last year after the Scheme and before the pandemic (in 2019 one in eight forego care while in 2021 just over one in ten). Then I considered whether this contradiction overturned the improvement after the Scheme. Having combined all indicators to capture *any* of the four outcomes (*All inclusive*), I refitted the model. The marginal probabilities are collected in table three (panel E). The same two patterns emerge: of marked reduction after the Scheme and of worse difference for families with older members.

Sensitivity analysis using 10% of total consumption shows the same two patterns of reduction after the Scheme and of higher risk among families with older members (panel F). However, the reduction is slight, having started from a low risk before the Scheme.

Remarkably, the higher risk to those families is of the same magnitude as obtained with all the other indicators using capacity to pay.

## Discussion

A well run health system delivers the goal of financial protection for all. Across three indicators of financial protection there is marked improvement after the Scheme for all families. Nevertheless, around one in eight families are still financially exposed and an increase in foregoing care has been masked, while families with older members remain at higher risk. This must be acknowledged alongside the Scheme’s well-recognised and well-qualified achievements.^2,5^

In ageing Asia, families with older members disproportionately incurred catastrophic payment including in Bhutan, China and Vietnam.^13,15,18,33^ In Vietnam in 2020, nearly one in eight is an older person, and among them more than one in eight foregoes care despite illness (:49).^34^ The analysis suggests that in Indonesia the Scheme has dampened this trend. Before and after the Scheme more families with older members (compared to other families) incurred catastrophic payment, but the difference is slight.

Foregone care compromises health systems, too. They can appear to perform well in financial protection while families carry on postponing treatment, a dissonance widely known yet often elided in middle and high income countries including Indonesia.^5,17^ Many health systems assessments forego analysing foregone care. Had this been conducted, the picture of financial protection would have been altered. For the first time the analysis shows that after the Scheme foregone care remained non-negligible and was associated with persistent social and spatial inequalities, including inequality across consumption quintiles. All this evidence on meeting the aims above suggests that the efficiency aim has been achieved at some cost of foregone care.

The key goal of financial protection needs to be continuously monitored, not least as the country transitions through a watershed launch of the national health insurance scheme. On the eve of its launch Indonesia’s President claimed that protecting older people was designed into the Scheme (<u>National Secretariat</u> 31 December 2013, accessed 31 Jan 2023).

However, in 2021 the joint report of the World Bank and WHO repeated the concern because families with older people have an ‘amplified risk’ of financial hardship.^34^ Indonesia has been able to dampen the amplified risk, though serious work remains. For instance, in response to a BBC journalist, Indonesia’s President suggested building more clinics in rural areas to alleviate persistent spatial inequalities cruelly exposed in the recent covid-19 pandemic (<u>BBC</u> <u>29 Oct 2021</u>, accessed 31 Jan 2023). This is echoed in Britain where the Chief Medical Officer in his annual report 2023 warns of rural and coastal ageing crisis (<u>BBC 10 November 2023</u>).

### Enhancing global initiatives by focusing on financial protection for older people

A focus of this study on families with older members is warranted because the UN’s Healthy Ageing Decade is nearly halfway, gaining traction along the way. Much can still be achieved in the global south where the population is ageing rapidly. Both the WHO and the Word Bank report that many of the older population live in the global south which puts an inordinate pressure on financing their health systems.^28,35^ The reports suggest a few strategies to respond, including deploying effective prevention, improving early diagnosis of hypertension and enhancing secondary prevention by using off-patent drugs to control blood pressure (:64).^28^ Since then our own work in Indonesia has delivered effective diagnosis and management of risk of cardiovascular disease in rural communities.^36^ Most importantly, this technology-based intervention has been proven to be cost effective, consequently adopted by a district government in East Java.^37,38^

Compared to before the scheme, the performance of the health system in 2021 is one marked with major achievements. These include increasing enrolment to cover nine in ten citizens of the third largest global south nation within a decade, registering the formal sector workers as well as the much larger informal sector, and spreading its operation across the archipelago which spans a distance similar to that from London (UK) to Doha (Qatar) or from Guayaquil (Ecuador) to Pernambuco (Brazil) or from Bamako (Mali) to Addis Ababa (Ethiopia) .^2,5^ Clearly, this cannot be achieved without challenges. Enrolment is not coverage, nor does coverage equate to quality or sustainability. The sustainable development goal of universal coverage by 2030 needs to recognise the important initiative of the UN Healthy Ageing Decade which also culminates at the same time. Both global initiatives must be cognizant of each other: efforts to secure universal health coverage can monitor what happens to families with older people, while interventions to deliver healthy ageing can monitor healthcare coverage of other age groups. Such keen recognition improves the likelihood of achieving sustainable and inclusive development goals.

### From healthcare back to health

The focus on monitoring financial protection whenever a person seeks care to recover health may struck critics as misplaced. After all, health is much more than healthcare. Health systems are often poised to ‘diagnose and treat’ rather than ‘predict and prevent’.^39^ But in middle income countries like Indonesia a focus on financial protection rightly holds its place because of the considerable number involved – in 2035 the older population of Indonesia is two thirds of the total UK population. Moreover, some conditions that demand healthcare arise from outside shocks such as climate change, whose local manifestations are difficult to predict or prevent (e.g. from heat stroke danger for older people). Diagnose/treat or predict/prevent are of course false dichotomies. Well-functioning health systems do both. In rural Indonesia our intervention using technology in the hands of community volunteers, reducing the risk of cerebrovascular events such as stroke, shows that it can be done in a way that also prevent risks turning to hazardous events.^36^

The retrospective observations used in this work put an important limit to inference – so there is no causal interpretation possible. Similarly, self-report of budget items is used, hence subjective errors may be present. Another limitation arises from lack of information on transportation costs to health facilities, a budget item found important elsewhere.^15^ Lastly, the year 2021 is the year of the pandemic, a unique event, one hopes. Nevertheless, all these observations are used regularly in recent assessments of Indonesia’s health system performance. These limits are more than balanced by the features of this work. It collated data from various sources and from times before and after the Scheme covering the geographic span of the archipelago. This strengthens the results obtained here as lessons for other global south nations undergoing rapid ageing. Another strength results from the modelling which conforms with the two levels of observations (families residing in districts). This has enabled an assessment of the balance between national (the *Jaminan Kesehatan Nasional* Scheme) and local support to financial protection, likely to be relevant to other countries where fiscal decentralisation is practised. Further research can progress from here including measuring foregone care as an integral indicator of financial protection.

To achieve universal health coverage whilst delivering healthy ageing, the monitoring of financial protection for all families especially those with older members is key. In Indonesia, while acknowledging the achievements, evidence has reinforced the need to remove the persistent social and spatial inequalities which mar the largest single-payer national health insurance scheme.

## Competing interest

I declare that I have no competing interests in conducting this investigation.

## STROBE statement

I have consulted the STROBE check list while writing this manuscript.

## Data Availability

This study uses data available to the public from the Central Bureau of Statistics Indonesia. https://sirusa.bps.go.id

## Notes

### Competing Interest Statement

The authors have declared no competing interest.

### Funding Statement

This study did not receive any funding.

